# CLIENT SATISFACTION ON CERVICAL CANCER SCEENING AND ITS ASSOCIATED FACTORS AMONG SCREENED WOMEN IN NORTH WOLLO PUBLIC HOSPITALS, ETHIOPIA

**DOI:** 10.1101/2024.06.16.24308997

**Authors:** Betelhem Ejigu, Tenagnwork Dilnesa, Toyba Ebrahim, Chaile Mulugeta, Belay Susu, Tadele Emagneh

## Abstract

**Background:** Cervical cancer is one of the very few cancers where pre-cancer lasts many years before becoming invasive cancer providing an opportunity for detection and treatment. It is leading cause cancer death where less attention is given for screening programs in sub-Saharan Africa. For this satisfaction of cervical cancer screening have a role in increasing utilization and quality of service

**Methods:** A facility based cross-sectional study design was used to assess client satisfaction and its associated factor towards cervical cancer screening. Data was collected from 397 samples using a face to face interview in women who take cervical screening test in the selected health facilities. Data was entered to Epi info version 4.6 and bivariable and multivariable logistic analysis was done using SPSS version 25 and p value <0.05 was considered to be significant factors associated with cervical cancer screening service

**Result:** from 397 interviewed women, 59% (236) were satisfied with cervical cancer screening services. One-hundred fourty five (36.5%) women had good comprehensive knowledge of cervical cancer screening. Satisfaction with cervical cancer screening service was statistically associated with Being house wife (AOR= 0.42(0.19-0.95), who attend Primary and secondary secondary (AOR =0.35(0.12-0.99) waiting time 31-60 min (AOR=0.14(0.06-0.33) and who went 1-5 km to the facility (AOR=0.41(0.19-0.90)

**Conclusion:** The study findings indicated that greater than half of women were satisfied with cervical cancer screening service in North wollo public hospitals. Less than half of women had good knowledge about cervical cancer. The study revealed occupational status, educational status, waiting time, and distance to the facility were found to be significantly associated with cervical cancer screening satisfaction

## Introduction

Cervical cancer is one of the very few cancers where a pre-cancer lasts many years before becoming invasive cancer providing an opportunity for detection and treatment(1). Cervical cancer screening is the systematic application of a test to identify cervical abnormalities in an asymptomatic population. Women targeted for screening may feel perfectly healthy and see no reason to visit health facilities(2).

There are various screening methods are pap smear test, visual inspection of the cervix with acetic acid or Lugol’s iodine: When vinegar is applied to abnormal cervical tissue, it temporarily turns white, and black or brown if iodine is applied whereas the normal retain its pink color Allowing the provider to make an immediate assessment of result. visual inspection of the cervix with acetic acid or Lugol’s iodine and are highly effective and affordable as a primary screening method for low resource settings because they do not rely on laboratory services (2).

Client satisfaction is an integral service-quality component that should be monitored closely by health service providers. A client is satisfied when his/her needs are met adequately when seeking healthcare services (3) while client satisfaction has multiple dimensions, key ones include communication, respect, client-provider relationship, provider characteristics, health facility and physical environment (4).

Patient satisfaction is considered a measure of the process of care according to Donabedian’s framework for health care evaluation (4). It is also related to the extent to which general health care needs and condition-specific needs are met(5). It can also be used to evaluate the quality of care, compare different health programs or systems, identify which aspects of a service need to be changed to improve patient satisfaction and also to assist health service providers to identify which patients are least likely to continue in a screening or therapeutic program (6).

Globally cervical cancer incidence rate is 3.1% and the most common cancer in 23 countries. It is the fourth most frequent cancer in women with an estimated 604 000 new cases and 342,000 cases of mortality globally in 2020(7,8).

In sub-Saharan Africa, cervical cancer is the leading cause of cancer death among females (9). It is estimated that 90% of cervical cancer deaths occurred in developing countries (10). A pooled prevalence of cervical cancer screening in Sub-Saharan Africa was 12.12% in 2021 (11).

In Ethiopia 20.9% of new case of cervical cancer occur in 2020 considering in both sex and all age (12).Cervical cancer is a second commonest disease of women in Ethiopia(13).The pooled prevalence of cervical cancer and precancerous lesion was found to be 15.7%, 15.6% (14,15). The pooled national level of cervical cancer screening among age-eligible women in Ethiopia and was 13.46% and 18.7% among HIV positive woman respectively (16,17).

Most (94.6%) of the women were satisfied with the screening service and 97% said they would recommend the test to others. The most common reasons for dissatisfaction with screening were discomfort during or after screening, long waiting time and failure to get treatment for other medical problems(18).

A research conducted in Addis Ababa confirmed that approximately half of the participants were not happy with the overall view and treatment approach that suggests acceptance of cervical cancer screening and treatment service among the study population. It also reveals lack of education and cervical cancer screening and care program awareness plan are the reasons for poor utilization of cervical cancer screening (19).

A national cancer control plan has organized and implemented from 2015 to 2020 by Ethiopia Federal Minister of Health (FMOH) which primarily targeted 30 to 49 years-old and high-risk groups of women of the screening and treatment for cervical pre cancer into 800 health facilities (one health facility per district) According to Tikur Anbessa Specialized Hospital, about 80% of reported cases of cancer are diagnosed at advanced stages, which is difficult to treat the disease. This is largely due to the low awareness of cancer signs and symptoms, inadequate screening and early detection and treatment services, inadequate diagnostic facilities and poorly structured referral(20).

Assessing patient satisfaction towards a service is clinically relevant, as satisfied clients are more likely to continue treatment if visual inspection of the cervix with acetic acid tested positive and recommend to take the screening to relatives and friends which can decrease cervical cancer mortality from early treatment of the disease and increase utilization of screening program and in turn decrease the incidence of cervical cancer(21).

There are few studies regarding satisfaction of cervical cancer screening so, determining clients level of satisfaction and factors that affect the satisfaction will help to increase utilization of the service and quality of service(21,22). Hence, this study primarily aimed to examine women’s level of satisfaction with cervical cancer screening services and factors affecting it.

## Methods

### Study settings and period

The study was done in North Wollo zone public hospitals providing cervical cancer screening service. North Wollo Zone, is one of 13 zones of the Amhara Region. There are 6 hospitals, and 69 health center found in this zone.

Woldia is capital city of North Wollo located 521 km far from Addis Ababa which is the capital city of Ethiopia and 360 km far from bahirdar. Cervical cancer screening service is available in all the six hospitals. The six hospitals are Wolidia comprehensive specialized hospital, lalibela general hospital, kobo primary hospital, Mersa primary hospital, shedehomekit hospital, and kone hospital primary hospital. The study was conducted was from November to December, 2023

### Study Design and participant selection

A facility based cross sectional study design was used and All women who received cervical cancer screening in North wollo public hospitals during study period were eligible Women who cannot communicate due to speech impairment were excluded from the study

### Data collection tool

Data was collected using face to face interview during their exit from services after obtaining their consent. To avoid social acceptability, bias the data collectors was not directly involved in giving cervical cancer screening service. The interviewed women were asked to give their honest response as confidentiality of the data will be kept Data was collected using a structured questionnaire which comprises sections of: women’s soico demographic characteristics, knowledge of cervical cancer screening and satisfaction with services regarding structure of area, service provision, privacy and autonomy which include 20 items with Likert scale. The questionnaires were adapted through review of relevant literatures the questionnaire were originally prepared in English and then translated to local languages Amharic and retranslated to English to check the consistency of questions. six midwives with BSc degree collected the data and another two midwives were supervisor

### Data processing and analysis

To assure data quality, completeness and consistency of data was checked, 5% pretest was done in Dessie Hospital before the actual data collection period and the credibility and transferability of the questions was revised, edited with necessary modification. Training was given for the supervisor and data collectors focused on the objective of the study, interview skill and issues related to the confidentiality of the response. The reliability of questions was evaluated using Cronbach’s alpha. In this study the level of Cronbach’s alpha is 0.92 which indicates high level of internal consistency for satisfaction questions

Data was entered by using Epi-info version 4.6 and exported to SPSS version 25 for analysis after data are coded and checked. Bivariable and multivariable analysis of logistic regression was used to assess the association of independent and dependent variables and P-value < 0.25, 0.05 was used to determine the level of statistical significance respectively. Odds ratio with 95% confidence interval was used to measure the strength of association. Descriptive statistic was used to summarize the data and the findings was presented using Tables and charts Client satisfaction: was assessed by structured Likert scale questionnaires which have five response options very satisfied, slightly satisfied, neutral, slightly dissatisfied and very dissatisfied. Those who scored (23)

Knowledge questions was scored, as score of 1 given for correct answer and zero for incorrect and don’t know and the mean score was computed. Women who scored above the mean was considered as having good knowledge. Women who scored below mean was considered as having poor knowledge

### Ethical consideration

Ethical clearance was obtained from ethical review committee of Wollo University College of Medicine and Health Science and formal letter was written to each of the selected health facilities verbal consent was taken from participants and recorded with audio. The study did not include minors. The data collectors explained that the information collected would be kept confidential and would be used for research purpose only. The objectives and benefits of the study was explained to the respondents. Data was collected if the respondents agree to participate in the study

## RESULT

### Socio-Demographic characteristics

A total of 397 women participated in the study. Majority of women were in the age range of 30– 49 years (78.6%) The average age of women was 39 years (SD+/-7.8) More than half (70%) of women were married and more than one-third (53%) were housewife. More than one fourth (38%) of women cannot read and 31% of them attended primary and secondary school

One-third (33%) of women were screened for cervical cancer screening. from them one –fourth of women were screened for the last time before 6-24 months. More than half percentage (76%) of women was waiting to get service for less than 30 min and more than two –fourth of women had to go 1-5 km from their home to the facility to get cervical cancer screening service. More than one fourth of women (28%) were appointed to cervical cancer screening service Table 1

**Table 1:** socio demographic characteristics of women in North wollo public hospitals,Ethiopia,2023.

| Variables | Frequency | Percent |
| --- | --- | --- |
| Age |  |  |
| <29 | 45 | 11.3 |
| 30-49 | 312 | 78.6 |
| >50 | 40 | 10.1 |
| Marital status |  |  |
| single | 37 | 9.3 |
| married | 280 | 70.5 |
| divorced | 57 | 14.4 |
| windowed | 23 | 5.8 |
| Occupation |  |  |
| house wife | 211 | 53.1 |
| government employee | 83 | 20.9 |
| merchant | 53 | 13.4 |
| daily worker | 50 | 12.6 |
| Education |  |  |
| cannot read and write | 152 | 38.3 |
| can read and write | 61 | 15.4 |
| primary and secondary | 123 | 31.0 |
| degree and above | 61 | 15.4 |
| Have you ever screened for cervical cancer? |  |  |
| yes | 131 | 33 |
| no | 266 | 67 |
| When is the last time of screening? |  |  |
| >6 months | 28 | 7.1 |
| 6-24 months | 91 | 22.9 |
| >24 months | 12 | 3.0 |
| Waiting time to get the service |  |  |
| <30min | 304 | 76.6 |
| 31-60min | 51 | 12.8 |
| >60min | 42 | 10.6 |
| Distance to the health facility |  |  |
| less than 1 km | 74 | 18.6 |
| 1-5 km | 220 | 55.4 |
| >5 km | 103 | 25.9 |
| Do you have an appointment for screening? |  |  |
| yes | 114 | 28.7 |
| no | 283 | 71.3 |

### Client satisfaction towards cervical cancer screening service

#### Satisfaction related to the basis of the institution and its staff

As shown below in the table from study participants 160 (40.3%) strongly satisfied when 108 (27.2%) were slightly satisfied regarding easy access to the diagnostic center. Many of the respondents 226 (56.9%) were strongly satisfied with the condition of being handled by the guards and none of women were strongly dissatisfied More than one -third of respondents 186 (46.9%) were strongly satisfied about the condition at the reception when they came to the screening center. Regarding the hygiene level around service 154(38.8%) were slightly satisfied and 110(27.7%) were strongly satisfied Table 2

**Table 2:**
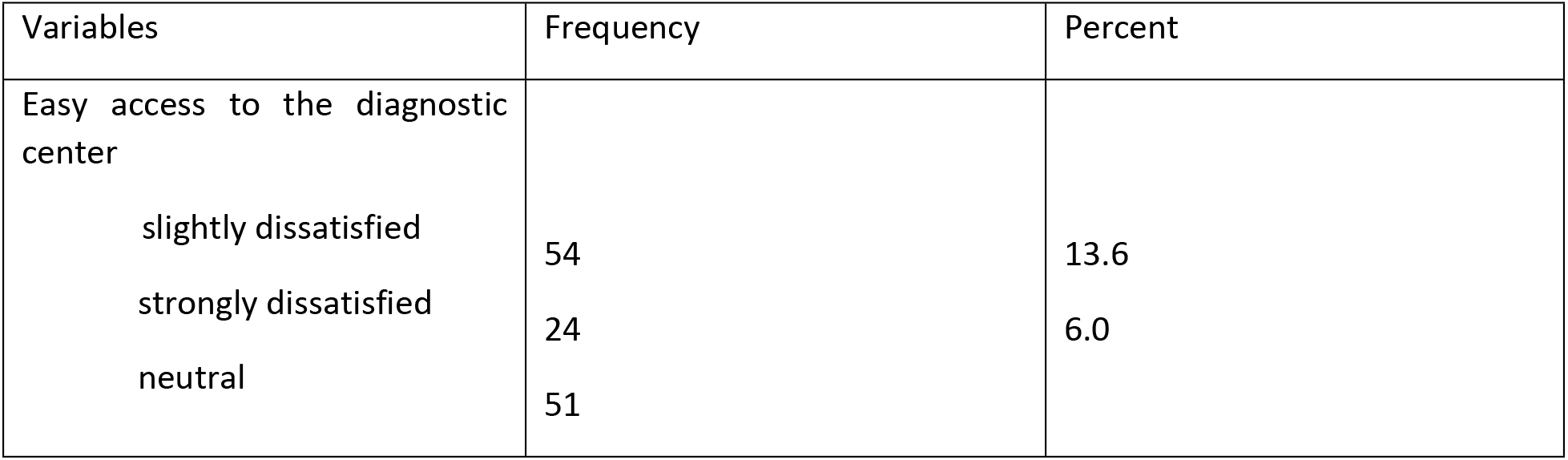
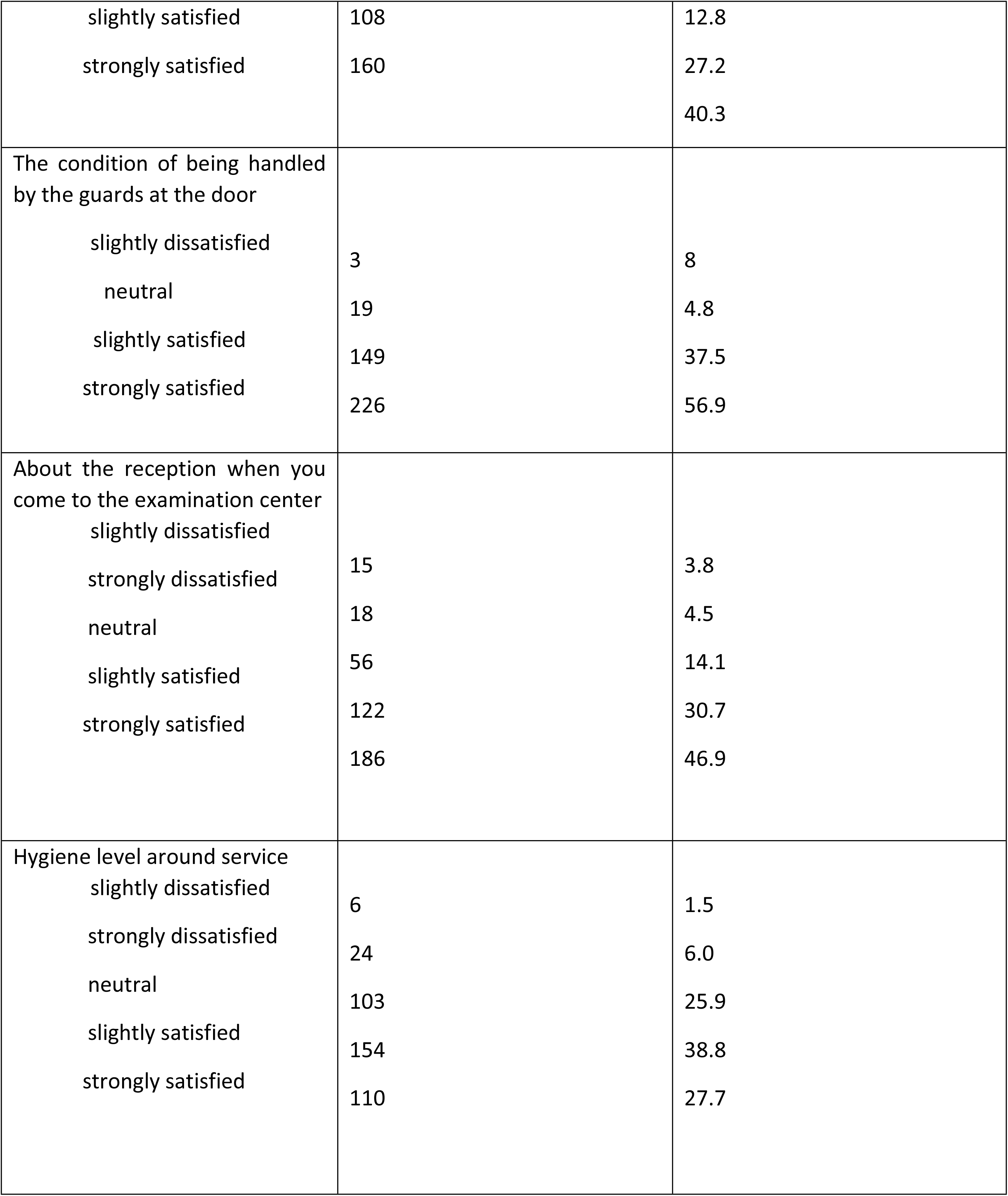
level of satisfaction related to basis of institution and its staff of screened women in North wollo public hospitals, Ethiopia,2023.

#### Satisfaction level related to service delivery process

Regarding the level of support for the diagnosis 162 (40.8%) of participants were strongly satisfied. Concerning the collaboration level of hospital workers around hospital environment 132 (33.2%) were strongly satisfied. In relation to the stage of explaining the diagnostic process 92 (23.2 %) were strongly satisfied and about the length of time before going to doctor 127 (32%) were strongly satisfied. Regarding the level of information received about diagnosis and treatment of cervical cancer screening more than one third 139(35%) of participants were strongly satisfied. About the level of service delivery of health professionals 145 (36.5%) of participants were strongly satisfied Table 3

**Table 3:**
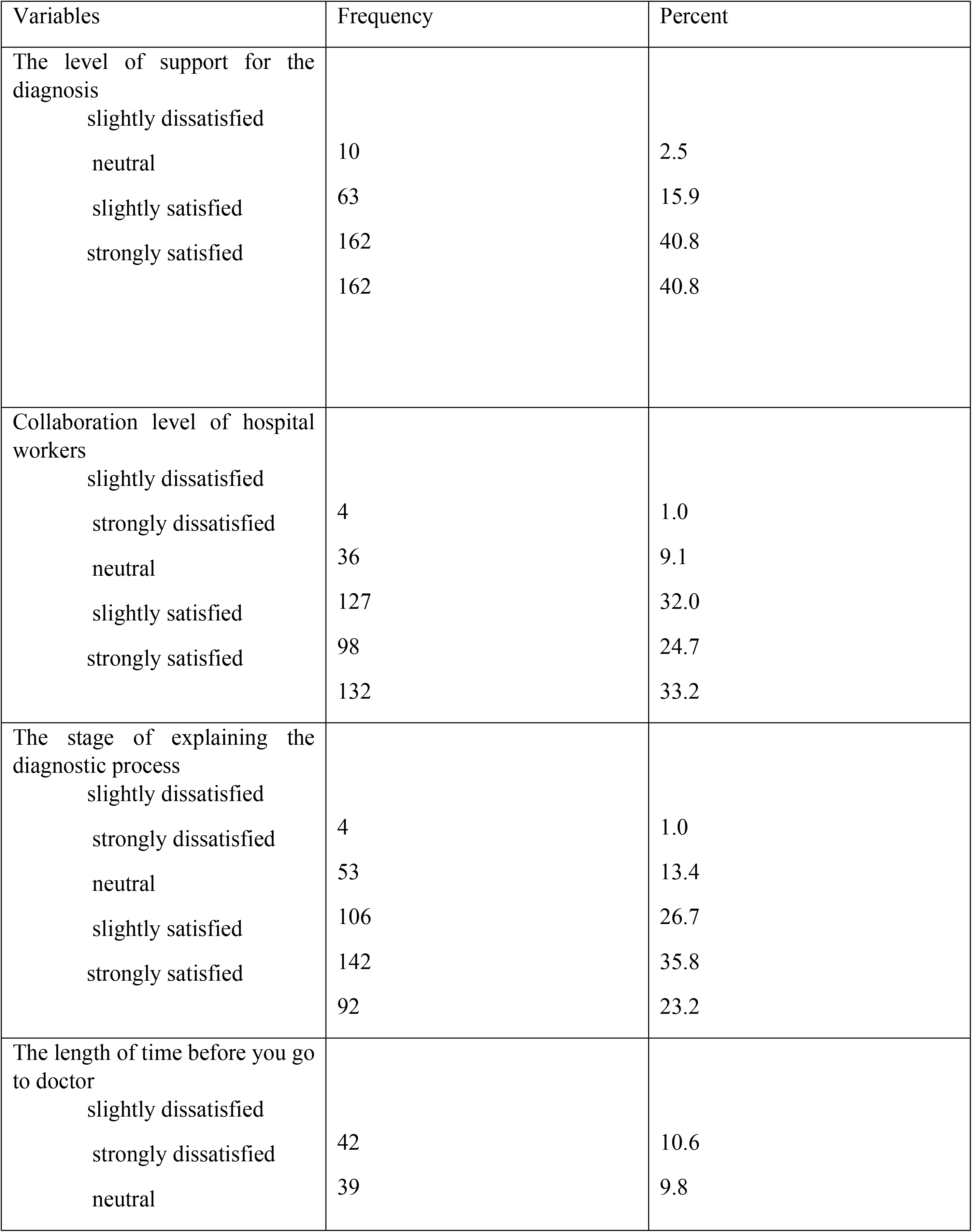
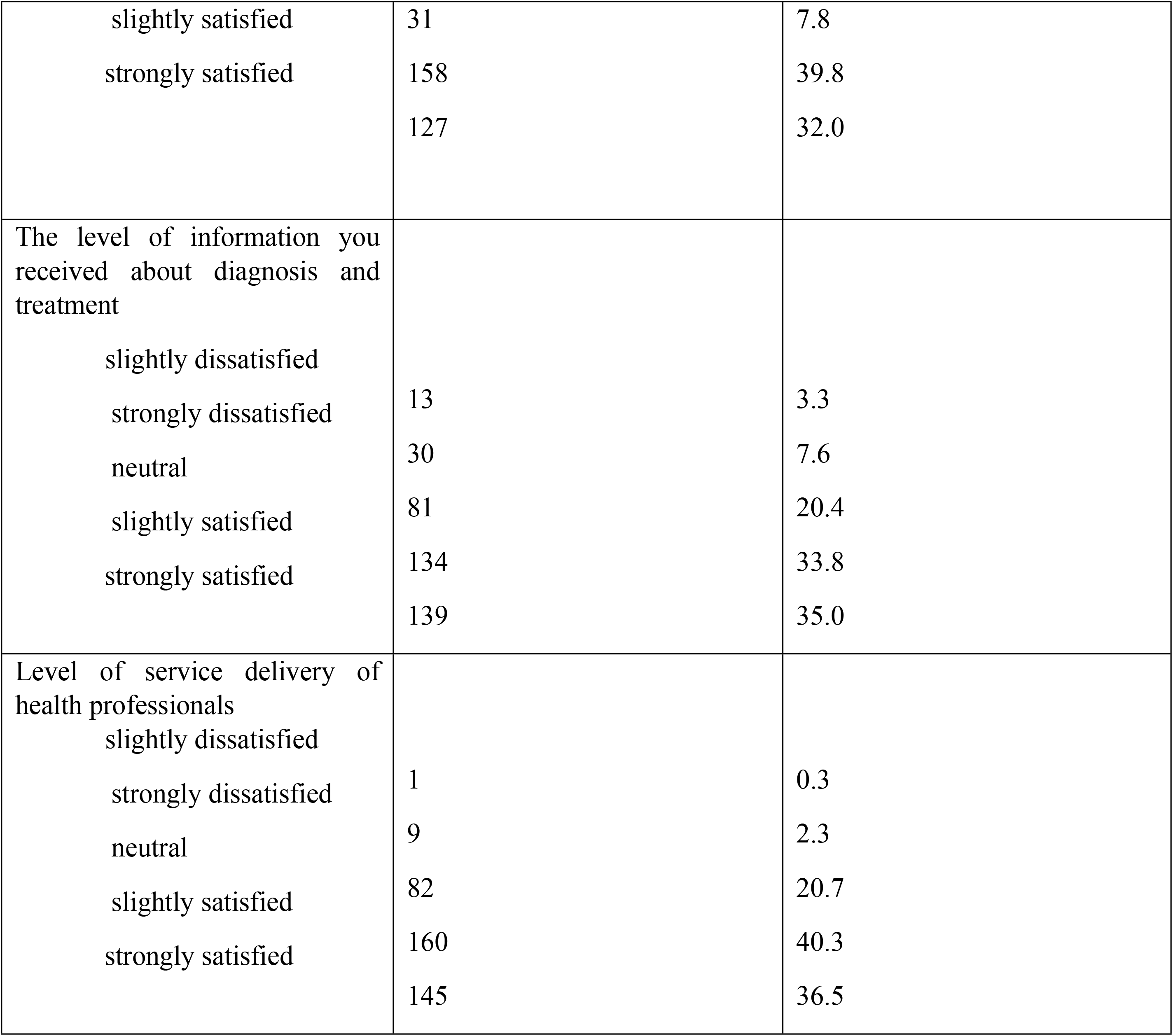
level of satisfaction related to service delivery process of screened women in North wollo public hospitals,Ethiopia,2023.

#### Satisfaction level of participants related to their privacy

Concerning the level of access to physician freely and privately 155(39%) of respondents were strongly satisfied and 181(45.6%) were slightly satisfied. In relation to the convenience of a room to express ideas without fear 160(40.3%) of women’s were strongly satisfied and 169(40.3%) of women’s were strongly satisfied about their personal freedom during examination. Regarding the Level of security of client’s personal information 162 (40.8%) was strongly satisfied and none of them were dissatisfied that indicates clients have trust on the physicians. About cleanness of examination instruments more than one fourth 124(31%) of women were strongly satisfied Table 4

**Table 4:**
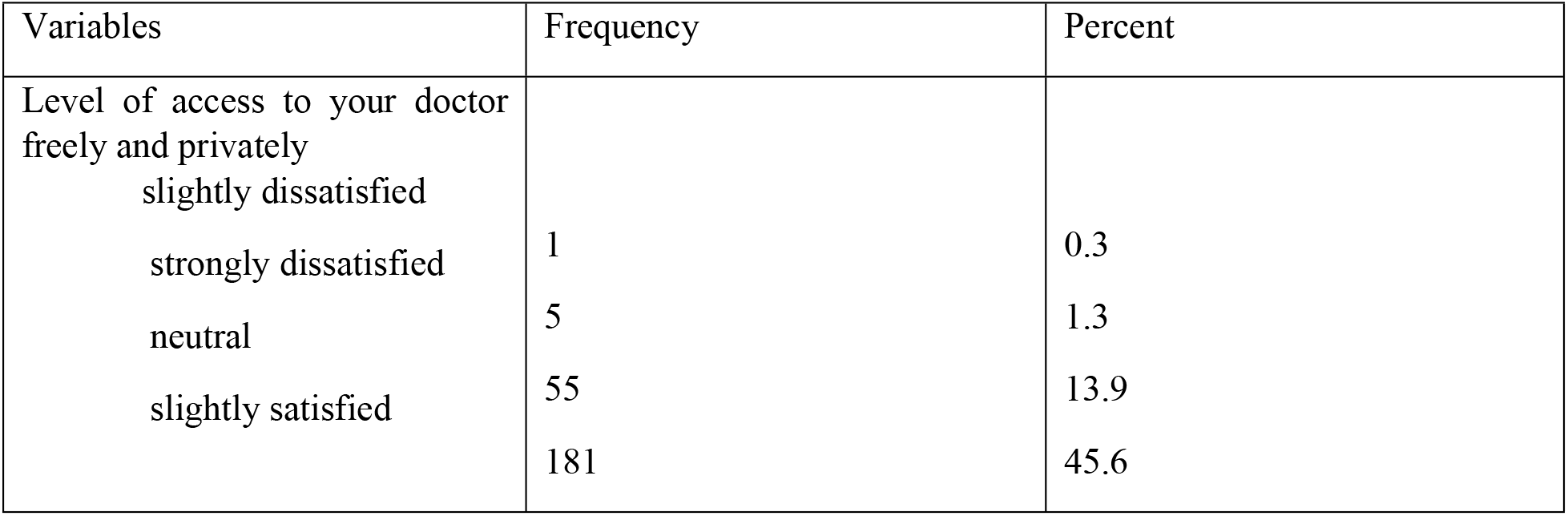
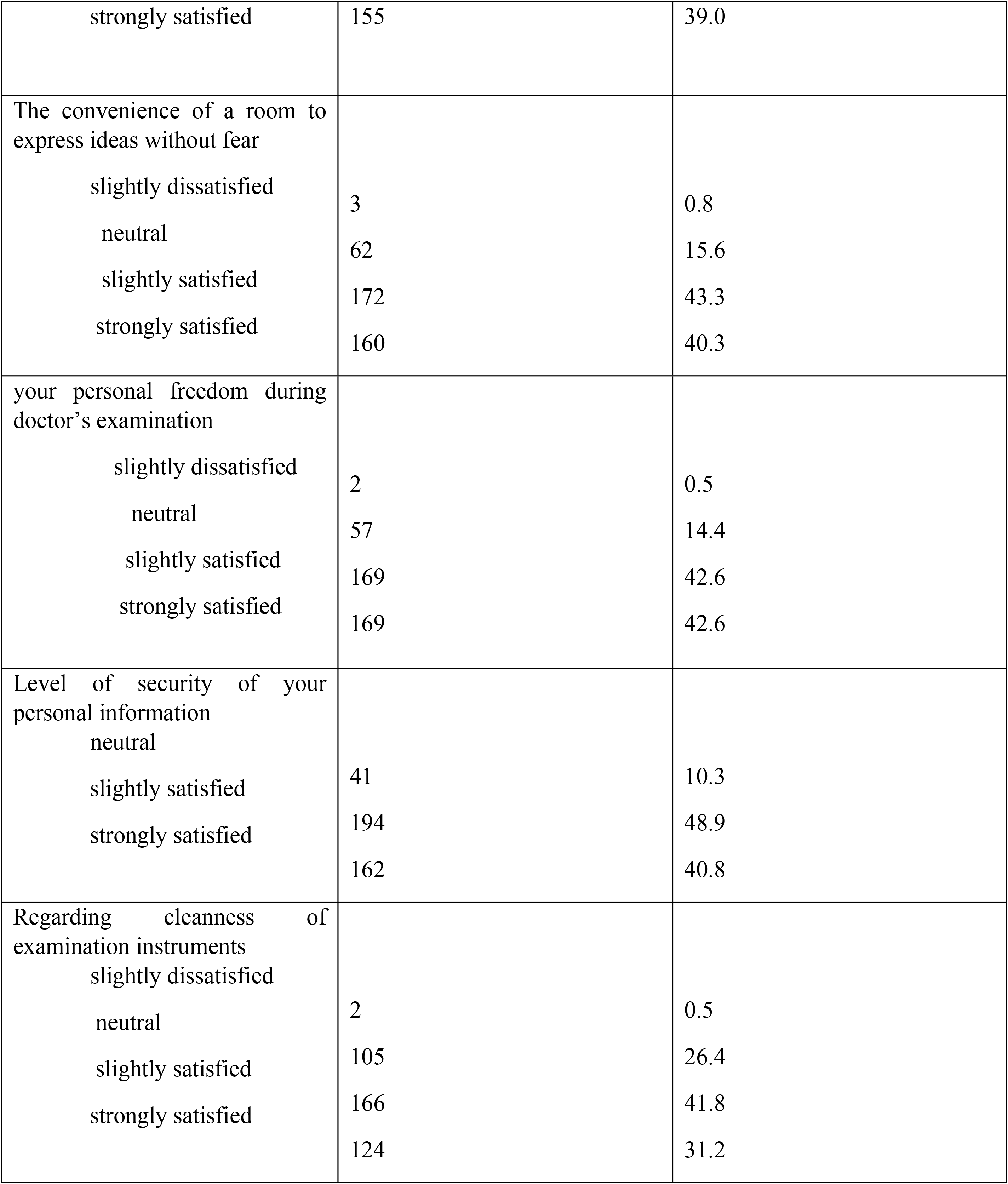
level of satisfaction related to privacy of screened women in North wollo public hospitals, Ethiopia,2023.

#### Satisfaction level related to autonomy

Regarding the right to take the test 112(28.2%) of women were strongly satisfied and the right to take treatment 115(29%) of them were strongly satisfied. Concerning the degree to which no one influence your decision122 (30%) of participants were strongly satisfied and about the opportunity to take time to think after service provision 121(30%) of women’s were strongly satisfied and 161(40%) were slightly satisfied Table 5

**Table 5:**
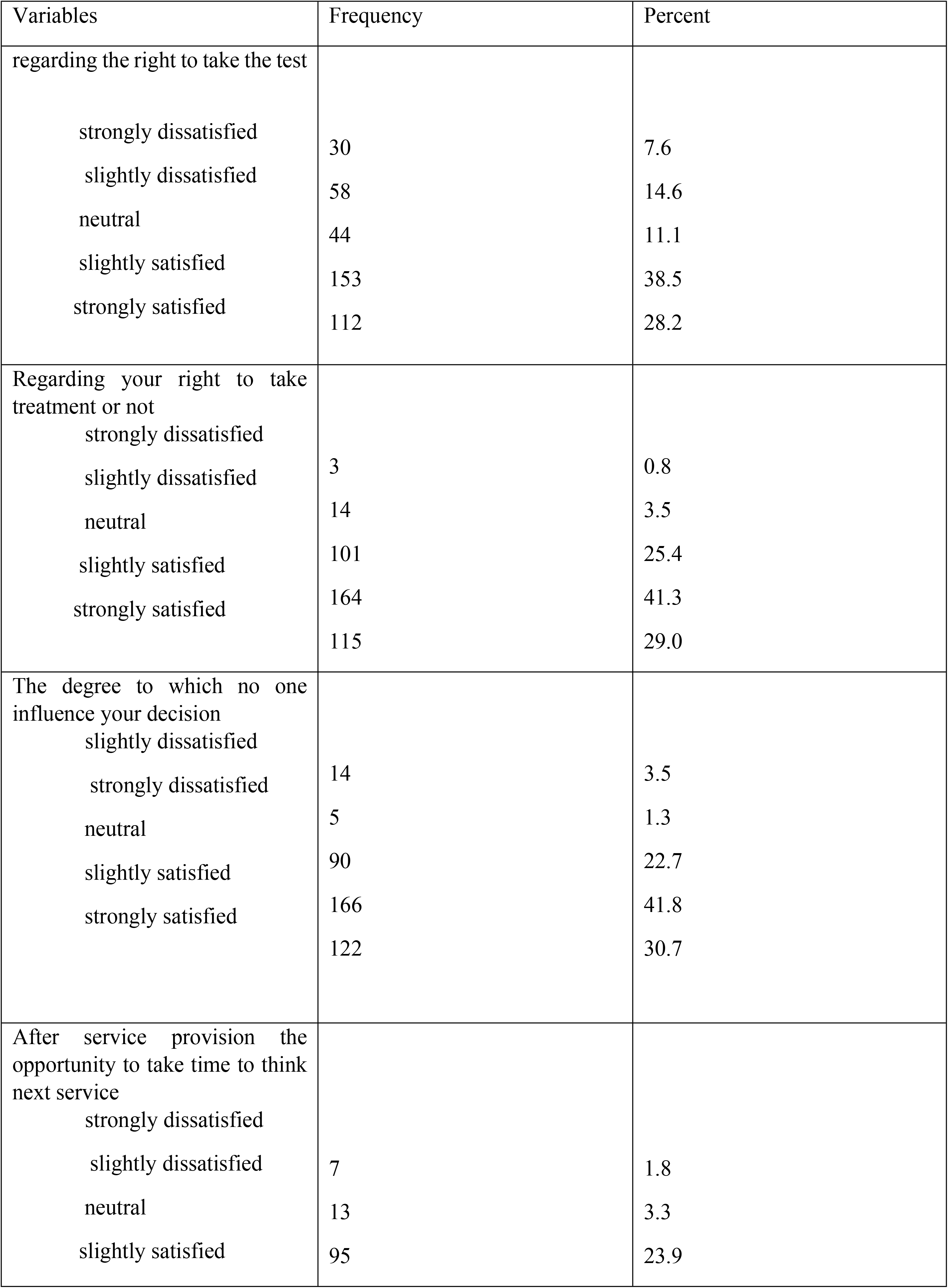
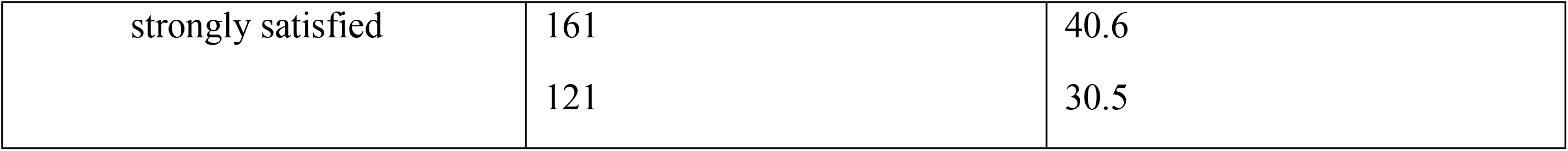
level of satisfaction related to autonomy of screened women in North Wollo public hospitals, Ethiopia,2023.

#### Overall satisfaction on cervical cancer screening service

From 397 women participated in the study 236(59%) of them were satisfied and 161(41%) of them were dissatisfied. satisfaction was measured on four major aspects which have different questions that are satisfaction on basis of institution and its staff, satisfaction on the service delivery, satisfaction related to privacy and autonomy. The mean score of satisfaction of cervical cancer service was 75.4 and those who scored above this level were considered satisfied and who scored below this level considered to be dissatisfied

#### Knowledge of cervical cancer

From all study participants, only 88(22.2%) of them had knowledge of risk factors of cervical cancer and 85 (21.4%) of listed multiple sexual partner, and 69(17.4%) listed early sexual initiation and less than one third listed exposing to HPV and smoking as risk factor. More than one fourth 127(32%) of participants had knowledge of symptoms of cervical cancer and 115 (29%) replied foul smelling vaginal discharge, 56(14.1%) pain during sex and 33(8.3%) as a symptom of cervical cancer screening. More than half percentage (54%) had known that cervical cancer can be cured if diagnosed earlier of participants Majority of participants 294 (74%) were aware of cervical cancer screening service availability. Overall knowledge of cervical cancer, 252(63.5%) of women had poor knowledge and 145(36.5%) had good knowledge of cervical cancer Table 6

**Table 6:** knowledge of cervical cancer among screened women at North wollo public hospitals,2023.

| Variables | Frequency | Percent |
| --- | --- | --- |
| Do you know the Risk factors of cervical cancer? |  |  |
| Yes | 88 | 22.2 |
| No | 309 | 77.8 |
| multiple sexual partner | 85 | 21.4 |
| exposing to HPV | 17 | 4.3 |
| early sexual initiation | 69 | 17.4 |
| smoking | 12 | 3 |
| Do you know the Symptom of cervical cancer? |  |  |
| Yes | 127 | 32 |
| No | 270 | 68 |
| Intermenstrual bleeding | 33 | 8.3 |
| pain during sex | 56 | 14.1 |
| foul smelling vaginal discharge | 115 | 29 |
| Do you Know cervical cancer can be cured if diagnosed early? |  |  |
| Yes | 227 | 57.2 |
| No | 170 | 42.8 |
| Do you Know availability of cervical cancer screening? |  |  |
| Yes | 294 | 74.1 |
| No | 103 | 25.9 |
| Overall knowledge |  |  |
| Poor | 252 | 63.5 |
| Good | 145 | 36.5 |

#### Factors associated with satisfaction of cervical cancer screening service

From eight variables entered in the bivariate logistic regression, educational status, occupation, distance from health facility, waiting time to get service, knowledge of cervical cancer and having appointment for the service were candidate variables for multiple variable logistic regression with P-value<0.25 In the multivariate logistic regression, occupational status, educational status, waiting time to get the service, distance to the health facility were significantly associated with cervical cancer screening services satisfaction with p value <0.05 Satisfaction with cervical cancer screening services was 74% (AOR=0.26; 95% CI (0.19-0.95) less likely among women who were housewives compared to daily workers. The odds of cervical cancer screening service satisfaction were four times more likely among Government workers than daily workers (AOR=4.55; 95% CI (1.46-14.1). women who have primary and secondary level of education was 65% (AOR=0.35; 95% CI (0.12-0.99) less likely to be satisfied than women who have degree and above level of educations.

The odds of cervical cancer screening service satisfaction were 86% (AOR =0.14;95% CI (0.06-0.33) less likely among women who wait 31-60 min to get the service than women who wait less than or equal to 30 min. women who went 1-5 km and 5 km from their home to the health facility were 59%, & 74% (AOR= 0.41; 95% CI (0.19-0.90) and (AOR=0.26; 95% CI (0.13-0.49) Less likely to be satisfied with cervical cancer screening service compared to women who went <1 km to the facility Table 7

**Table 7:** factors associated with client satisfaction of cervical cancer screenin among screened women in North Wollo public hospitals,Ethiopia,2023.

| Variables | Satisfied | Dissatisfied | COR(95% CI) | AOR(95% CI) |
| --- | --- | --- | --- | --- |
| Age |  |  |  |  |
| <29 | 25(10.6%) | 20(12.4%) | 1.48(0.61-3.56) |  |
| 30-49 | 185(78.4%) | 127(78.9) | 1.27(0.64-2.53) |  |
| >50 | 26(11%) | 14(16%) | 1 |  |
| Marital status |  |  |  |  |
| Single | 18(7.6%) | 19(11.8%) | 1.37(0.48-3.90) |  |
| Married | 174(73.4%) | 106(65.8%) | 0.79(0.33-1.87) |  |
| Divorced | 31(13.1%) | 26(16.1%) | 1.09(0.41-2.89) |  |
| Windowed | 13(5.5%) | 10(6.2%) | 1 |  |
| Occupation status |  |  |  |  |
| Housewife | 160(67.8%) | 51(31.7%) | 0.90(0.44-1.83) | 0.42(0.19-0.95) |
| Government employee | 16(6.8%) | 67(41.6%) | 11.91(5.17-27.4) * | ** |
|  | 23(9.7%) | 30(18.6%) | 3.71(1.61-8.54) * |  |
| Merchant | 37(15.7%) | 13(8.1%) | 1 | 4.55(1.46-14.1)<br>** |
| Daily worker |  |  |  | 2.09(0.806-5.45)<br>1 |
| Educational status |  |  |  |  |
| cannot read and write | 113(47.9%) | 39(24.2%) | 0.06(0.02-0.132) * | 0.41(0.13-1.28) |
| can read and write | 38(16.1%) | 23(14.3%) | 0.10(0.04-0.25) * | 0.31(0.09-1.09) |
| primary and secondary | 76(32.2%) | 47(29.2%) | 0.107(0.04-0.23) * | 0.35(0.12-0.99) |
| degree and above | 9(3.8%) | 52(32.3%) | 1 | **<br>1 |
| Waiting time |  |  |  |  |
| <30min | 218(92.4%) | 87(54%) | 1 |  |
| 31-60min | 6(2.5%) | 44(27.3%) | 0.16(0.07-0.32) * | 0.14(0.06-0.33) |
| >60min | 12(5.1%) | 30(18.6%) | 2.93(0.99-8.67)* | **<br>1.65(0.47-5.76) |
| Distance to the health facility |  |  |  |  |
| less than 1 km | 45(19.1%) | 29(18%) | 1 | 1 |
| 1-5 km | 152(64.4%) | 68(42.2%) | 0.39(0.21-0.72) * | 0.41(0.19-0.90) |
| >5 km | 39(16.5%) | 64(39.8%) | 0.27(0.16-0.44)* | **<br>0.26(0.13-0.49)<br>** |
| Appointment |  |  |  |  |
| Yes | 77(32.6%) | 37(23%) | 1.62(1.02-2.56) * | 0.73(0.40-1.33) |
| No | 159(67.4%) | 124(77%) | 1 | 1 |
| knowledge |  |  |  |  |
| Poor | 175(74.2%) | 77(47.8%) | 3.13(2.04-4.78) * | 1.09(0.57-2.09) |
| Good | 61(25.8%) | 84(52.2%) | 1 | 1 |

## Discussion

The study has shown that overall satisfaction with cervical cancer screening was 59%. maximum number 312(78.6%) of women screened with VIA were in the age of 30-49 which is according to ministry of health recommendation. In this study, occupational status, educational status, waiting time to get the service, and distance to the health facility were identified as factors that affect cervical cancer screening services. We found no evidence that women’s level of satisfaction with the cervical cancer screening services was associated with knowledge toward cervical cancer.

The overall cervical cancer screening service satisfaction in this study area was comparable to the study done in Addis Ababa and was slightly lower than study done in jimma(21,23).Another study in Malawi shows that 68.3% of women were very satisfied with the service they received which is much higher than in this study(24).

The difference may be due to satisfaction measurement questionnaires considered and study area difference. In this study more than half percentage of women were strongly satisfied with the condition being handled by the guards and least satisfied with waiting time to get the service

In the study area less than half of women were slightly satisfied with level of information received about diagnosis and treatment of cervical cancer screening which is much lower than study done in Jimma, Addis Ababa and Malawi(21,23,24) the finding indicates that there is health professional’s gap in explaining the information about diagnosis and treatment of cervical cancer screening service in this study

Less than half of women had Good Knowledge of about cervical cancer risk factor, symptom, curability and availability of screening service. This finding was lower than study conducted in Durame, Jimma, Dessie, Tigray, Wolayta, contrary the finding was higher than study done in Gondar(21,25–29).The variation may be related to knowledge measurement considered and study area difference

There is significant association between socio demographic factors (occupation, education, distance) and cervical cancer screening service. Government workers were four times more likely to be satisfied than daily workers. Occupational status was also found associated in other studies. Marital status and age were not associated with cervical cancer screening in this study which is also reported by other studies (21,24).

The study finding also indicated a significant association between service characteristics (waiting time to see a provider) and women’s satisfaction with cervical cancer screening services. Women who had waited 31-60 min to get the service were 86% less likely to be satisfied than who had waited 30 min and less. The finding was consistent with other studies (21,24).

Distance to the facility was significantly associated with cervical cancer screening service. Women who went 1-5 km were less likely to be satisfied than who went < 1km which is similarly reported in study done in Jimma contrary, distance to the health facility were not significantly associated in study of Malawi(21,24).

In our study, knowledge of cervical cancer and having appointment were associated with women’s satisfaction in the Univariate analysis. However, these factors did not show association in the multivariate analysis. In other studies knowledge of cervical cancer and having appointment were found to have significant association(21,24).

## Conclusion

The study findings indicated that greater than half of women of were satisfied with cervical cancer screening service in North wollo public hospitals. Less than half of women had good knowledge about cervical cancer risk factors; symptoms, curability, and availability of screening service. The study revealed that occupational status, educational status, waiting time to get the service, and distance to the facility were found to be significantly associated with satisfaction of cervical cancer screening service. Therefore, Health facilities need to create health education programs regarding cervical cancer knowledge and cervical cancer screening advantage, Health care providers need to available in working hours in screening room so that clients need not to wait to get the service.

**Figure 1:**
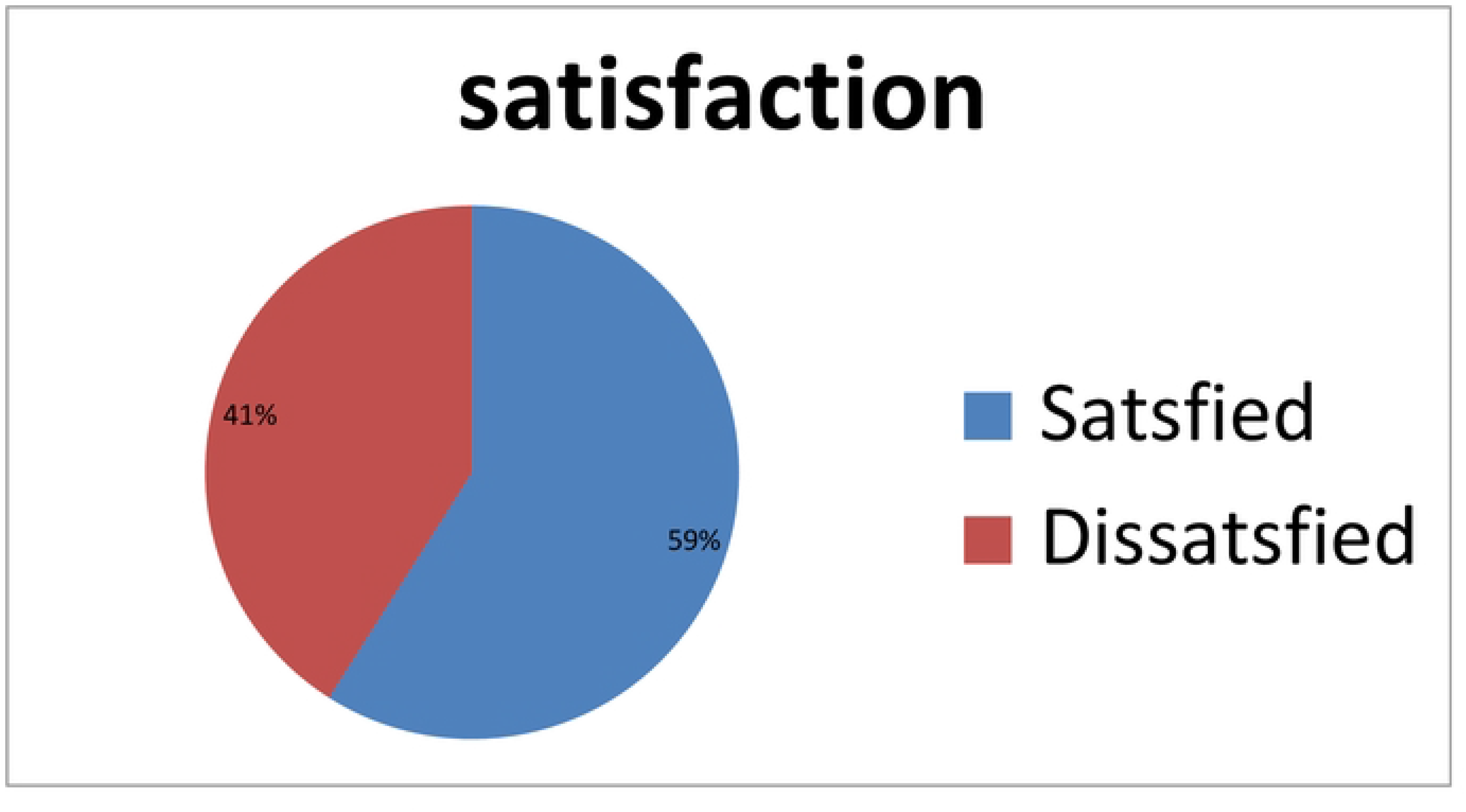
overall satisfaction of cervical cancer screening service among women screened women in North wollo public hospitals, Ethiopia,2023.

## Data Availability

NO

## Limitation of the study

The study was conducted in six public hospitals only which may limit generalizability of the findings to women who get the service in health centers. The study used only quantitative method to measure satisfaction of cervical cancer screening service which would be better if it includes both quantitative and qualitative method

## Acknowledgments

We acknowledge the different levels of administrative hierarchy and study participants.

## Author Contributions

All authors made a significant contribution to the work reported, whether that is in the conception, study design, execution, acquisition of data, analysis and interpretation, or in all these areas; took part in drafting, revising or critically reviewing the article; gave final approval of the version to be published; have agreed on the journal to which the article has been submitted; and agree to be accountable for all aspects of the work.

